# The recurrence of illness (ROI) index is a key factor in major depression that indicates increasing immune-linked neurotoxicity and vulnerability to suicidal behaviors

**DOI:** 10.1101/2024.04.11.24305652

**Authors:** Michael Maes, Ketsupar Jirakran, Asara Vasupanrajit, Mengqi Niu, Bo Zhou, Drozdstoj St. Stoyanov, Chavit Tunvirachaisakul

## Abstract

**Background:** The first author of this paper introduced new precision nomothetic models for a major depressive episode (MDD) which incorporate quantitative scores that measure recurrence of illness (ROI).

**Objective:** To explore the connections between ROI and biomarkers related to an activated immune network, immune-linked neurotoxicity (INT), and a combined INT and atherogenicity index (METAMMUNE).

**Methods:** The study involved 67 healthy controls and 66 outpatient MDD (OMDD) participants. We utilized a Multiplex method to measure 48 cytokines, and developed INT and METAMMUNE composite scores. The measurements included triglycerides, free cholesterol, LDL and HDL-cholesterol, apolipoprotein (Apo)A1 and ApoB.

**Results:** A ROI index was successfully created by extracting a validated principal component, from the number of physician-rated depressive episodes, the frequency of lifetime suicidal ideation and attempts. Adverse childhood experiences accounted for 20-22% of the variance in ROI. The latter was significantly associated with INT and METAMMUNE indices, neuroticism, lifetime and current suicidal behaviors, and the phenome (p<0.001). Our analysis revealed that a significant portion (55.1%) of the variance in the OMDD phenome, which includes current suicidal behaviors, anxiety, and depression, can be accounted for by the regression on INT, ROI, and emotional neglect and abuse. A validated latent construct was successfully extracted from the three ROI components, INT and METAMMUNE indices.

**Conclusions:** ROI and associated immune-metabolic biomarkers are indicators of a shared underlying concept, specifically a ROI-neuroimmune-metabolic pathway phenotype. ROI is a crucial indicator of the rising immune-metabolic abnormalities and heightened susceptibility to suicidal tendencies and recurrence of illness.

## Introduction

In recent publications, the first author of this paper developed novel precision models for a major depressive episode (MDD) using the precision nomothetic approach [1–9]. The author utilized a theoretical framework to construct a clinical precision nomothetic model that connects clinical causal factors (such as a family history of mood and substance use disorders) and adverse childhood experiences (ACEs) with the lifetime and current phenome of depression [1,3,4,5,7]. The current phenome of MDD was understood as a factor (latent vector) derived from various scales used by psychiatrists. These scales assess the severity of depression, anxiety, chronic fatigue, self-rated severity of depression and anxiety, suicidal behaviors (SB), and self-rated health-related quality of life (HR-QoL). Therefore, the MDD phenome is conceptualized as a quantitative score based on a multitude of relevant ratings rather than a binary variable indicating the presence or absence of MDD [1,2,4,5,6,7,8,9]. As mentioned by other authors, the binary diagnosis based on DSM criteria should not be used in depression research or clinical practice. In addition, the concept of lifetime phenome was derived from neuroticism scores, the presence of lifetime anxiety disorders, and dysthymia [7]. In previous studies, the author of this paper found a strong correlation between the lifetime phenome and current phenome scores [7].

Research has found that both a positive family history and ACEs, such as physical and emotional neglect, can be predictors of the MDD phenome [3,4,5,7,8]. Additionally, it was discovered that some of these effects were mediated by the lifetime phenome of MDD [7]. Those models were built using partial least squares (PLS)-SEM analysis, which integrates factor analysis and multiple regression. At least 5,000 bootstrap samples were utilized in a multistep mediation model. After careful examination of the model fit criteria, these models were validated and found to have strong replicability [1,2,7].

Crucially, these clinical precision models also included an index of the recurrence of depressive episodes and SBs [1,3,4,5,6,7,8]. It is widely recognized that mood disorders, such as MDD, are progressive in nature. As the number of episodes increases, there is a corresponding increase in the severity of the current phenomenon and a decline in HR-QoL [3,4,7]. Various authors have proposed staging models that classify mood disorders into distinct stages [10–12]. The stages were identified based on different risk factors, including a history of mental illness in the family, patterns of early symptoms, recurrence of episodes, treatment resistance, and the level of functioning during non-episodic periods [10–12]. However, these staging models were theoretical concepts and did not incorporate predictive mathematical algorithms based on real patient data. In contrast, the primary author employed machine learning techniques, specifically factor and principal component (PC) analysis, to calculate a novel staging index known as recurrence of illness (ROI) [3,4,7]. This ROI index was developed as the first PC or factor extracted from the total number of depressive episodes experienced throughout one’s lifetime, along with the frequency of lifetime suicidal thoughts (SI) and suicide attempts (SA). Significantly, the impact of ACEs on the present phenome was not only influenced by the lifetime phenome, but also by ROI. It is worth noting that a significant portion (68.5%) of the variance in the current phenome can be attributed to ROI and lifetime phenome, as indicated by Maes et al. [7]. In addition, the combination of ROI and lifetime phenome could be seen as a single factor, suggesting that both concepts are reflective manifestations of the lifetime trajectory of patients with mood disorders [7]. The presence of certain lifetime epochs, such as ACEs, recurrent acute episodes and SB (ROI), neuroticism, anxiety disorders and dysthymia, play a significant role in shaping the characteristics of the current depressive phenome.

Extensive evidence suggests that MDD is a neuroimmune disorder, where immune pathways intersect with metabolic pathways, leading to heightened immune-linked neurotoxicity (INT) [5,8,9,13,14,15,16,17]. The research team led by the primary author of the study has successfully developed a variety of precision nomothetic models to analyze the acute phase of inpatient MDD (IMDD). These models incorporate factors such as ACEs, ROI, the phenome, all linked to the intersections of neuroimmune and metabolic pathways [1,2,5,7,8,9]. The latter encompass cytokine networks, T cell activation markers and T regulatory cells, heightened atherogenicity, elevated presence of Gram-negative bacteria, irregularities in the gut microbiome, oxidative stress, antioxidant levels, and neurotrophic pathways. Within the various models, ROI emerged as a crucial element, showing a strong correlation with biomarker pathways and ultimately impacting the phenome of acute and severe IMDD. In certain cases, a single factor was derived from the three ROI components and INT networks, resulting in the formation of a ROI-INT pathway phenotype [5].

However, it is still unclear whether the connections between ACEs, ROI, the lifetime phenome, INT and metabolic pathways, and the current phenome can be established in a representative sample of outpatients with MDD (OMDD) who have varying levels of ROI and mild to severe MDD. In a recent study, the research team led by the first author examined the neuroimmune and metabolic profile of OMDD. They found that OMDD is associated with activated immune pathways, INT, and atherogenicity and their interactions [18]. One interesting discovery in patients with OMDD is that a significant portion of the variation in the severity of the current OMDD phenome (31.4%) can be reliably predicted by the impact of INT and the interaction between INT and the metabolic syndrome (MetS) [18].

Therefore, this study aims to investigate the role of ROI in OMDD. Specifically, we will explore the potential connections between ROI and both the lifetime and current phenome. Additionally, we will examine how ROI may mediate the effects of ACEs on the lifetime and current phenome. Furthermore, we will analyze the associations between ROI and INT/metabolic biomarkers of OMDD. Another question is whether the self-rated number of depressive episodes is a valid index to reflect episode number.

## Methods and Participants

### Participants

This study included a group of sixty-seven healthy controls and another group of 66 patients diagnosed with OMDD. OMDD patients were enrolled as outpatients at the Department of Psychiatry in King Chulalongkorn Memorial Hospital in Bangkok, Thailand. The outpatients received a diagnosis of MDD based on the criteria specified in the DSM-5 [19]. The OMDD sample we have collected accurately reflects the population of OMDD in Bangkok. This is evident from the wide range of the number of depressive episodes, which spans from 1 to 20 episodes, and the wide range of scores on the Hamilton Depression Rating Scale (HDRS) scores which varied from 7 to 33. In addition, the sample included severe and mild depression, and encompassed both non-responders and partial remitters to treatment.

The control group was recruited through verbal communication within the catchment area, specifically in Bangkok, Thailand. Individuals in good health who participated in the study included personnel, relatives of staff colleagues, and acquaintances of individuals diagnosed with OMDD. To analyze the influence of MetS on immune profiles, we selected approximately half of the controls and OMDD samples who had been diagnosed with MetS. A total of sixty-four participants were found to have MetS, while sixty-nine did not exhibit the condition. Out of the total of sixty-seven controls, a significant number of thirty-three were found to have MetS. Similarly, among the 66 patients with OMDD, a considerable proportion of thirty-one were diagnosed with MetS.

The exclusion criteria that apply to both patients and controls are: a) neurological disorders, such as epilepsy, stroke, Parkinson’s and Alzheimer’s disease, multiple sclerosis, and brain tumors; b) DSM-5 axis-2 disorders, including borderline personality disorder and antisocial personality disorder; c) other axis-1 psychiatric disorders, such as autism spectrum disorders, substance use disorders, bipolar disorder, psycho-organic syndromes; schizophrenia, and schizo-affective psychosis; and d) pregnant or lactating women. Furthermore, individuals who met the criteria for the control group and had a positive familial history in first-degree relatives of suicide, depression, mania, substance use disorders, or suffered from current and/or lifetime MDD and dysthymia were excluded from the study.

Prior to their participation in the study, all participants willingly and knowingly gave their written informed consent. The research was conducted in accordance with ethical standards that are recognized globally and in Thailand, as well as in compliance with privacy laws. The inquiry received authorization (#445/63) from the Institutional Review Board of the Faculty of Medicine at Chulalongkorn University in Bangkok, Thailand.

### Clinical assessments

By utilizing a structured questionnaire and conducting interviews with both patients and controls, we collected sociodemographic and clinical data. The structured interview included a set of demographic questions, number of depressive episodes, family history of mental health issues in first-degree family members, the applicant’s medical history and use of psychotropic and medical medications. The assessment of the number of lifetime depressive episodes involved two methods: self-rating and extraction from the patient’s files at the outpatient department (OPD). Most patients received treatment at our OPD from the beginning of their disorder. For a minority of patients, we recorded the number of episodes they experienced prior to visiting our outpatient department through hetero anamnesis. Therefore, an accurate approximation of the number of episodes was acquired using the OPD files. The severity of MDD was assessed by utilizing the Beck Depression Inventory II (BDI-II) and the Hamilton Rating Scale for Depression (HAMD) [20,21]. Anxiety severity was scored using the state version of the State-Trait Anxiety Inventory (STAI) [22]. The diagnosis of MDD involved the use of the Mini International Neuropsychiatric Interview (M.I.N.I.), in a validated Thai translation [23], along with the DSM-5 criteria. In addition, the MINI was utilized for diagnosing other axis-1 conditions and for excluding other diagnosis.

A Thai translation of the ACE Questionnaire [24] was used to conduct the assessment of ACEs. The survey consists of twenty-eight items that relate to traumatic experiences in childhood and covers ten different areas. For the purposes of this research, we utilized the raw summed scores on various factors including emotional abuse, physical abuse, emotional and physical neglect, sexual abuse, domestic violence, substance abuse within the household, mental illness within the household, parental divorce, and a criminal household member. The current study also included the overall number of ACEs and the sum of three ACE subdomains, namely emotional abuse + emotional neglect + sex abuse (labeled as 3ACEs). Using the C-SSRS [25], we assessed the frequency, severity, intensity of suicidal behaviors (SB) including suicidal ideation (SI) and attempts (SA). The symptoms that were present during the month before admission into the study were referred to as “current” (current_SI or current_SA), while the symptoms that occurred earlier in a person’s lifetime were referred to as “lifetime” (LT_SI and LT_SA). In the current study we used two C-SSRS items, namely frequency of LT_SI and LT_SA. Furthermore, as explained previously, we extracted one factor from the current_SI and current_SA items (labeled current_SB). To assess major personality traits, we used the Thai translation of the Big Five Inventory (BFI) [26,27]. In this study, the researchers utilized the neuroticism scores, which were identified as a significant factor in the LT phenome of MDD [28]. The recurrence of illness (ROI) index was computed by utilizing the number of depressive episodes, LT_SI, and LT_SA items to derive the first principal component. The associations with other biomarker and clinical data were investigated in the total study sample and in the restricted group of MDD patients. In addition, we also investigated an adjusted ROI index (labeled adjusted ROI) which used the number of episodes – 1 (thus considering the first episodes as zero episodes). The latter index was used to minimize the effect of the diagnosis (MDD versus HC). Moreover, the results were also checked after transforming the ROI scores in fractional rank and log10 transformation. The severity of the LT phenome was conceptualized as the first PC extracted from neuroticism, number of LT anxiety disorders, and dysthymia [7]. The severity of the current OMDD phenomene (labeled as current_phenome) was determined as the first principal component derived from the HAMD, STAI, BDI and current_SB scores [7]. As mentioned in the Statistics section, each of these PCs met the prespecified quality criteria with great rigor.

The diagnostic criteria outlined in the DSM-5 were used to diagnose tobacco use disorder (TUD). The BMI was determined by dividing the weight (in kilograms) by the square of the height (in meters). MetS was defined based on the criteria outlined in the 2009 Joint Scientific Statement of the American Heart Association/National Heart, Lung, and Blood Institute [29]. It requires the presence of three or more of the following components: a) a low HDL-cholesterol levels, b) a high triglyceride level, c) an increased waist circumference or BMI, d) elevated fasting glucose levels or diabetes diagnosis, and e) high blood pressure.

### Analyses

During the designated time frame of 8:00 and 9:00 a.m., we collected a volume of twenty-five milliliters of fasting venous blood from every participant in the study. This was done by utilizing a serum tube and a disposable pipette. Next, the blood was spun at a rapid speed of 35,000 revolutions per minute to separate its components. The resulting serum was carefully collected and stored in Eppendorf tubes. These containers were then kept at a freezing temperature of −80 degrees Celsius until they were thawed for the biomarker assays. A total of forty-eight cytokines, chemokines, and growth factors were examined to determine their fluorescence intensities (FI) using the Bio-Plex Multiplex Immunoassay reagent provided by Bio-Rad Laboratories Inc., located in Hercules, USA [30]. As explained in the latter study, we successfully identified the first principal component derived from 10 differentially expressed and upregulated cytokines/chemokines/growth factors (labeled immune network index). Most importantly, this PC showed high loadings (>0.60) on 10 cytokines/chemokines/growth factors, and protein-protein interaction (PPI) analysis showed that these 10 immune compounds formed a tight PPI network [30]. The 10 upregulated components are: interleukin-4 (IL-4), IL-9, IL-17, CCL4, CCL5, CXCL12, PDGF (platelet-derived growth factor), tumor necrosis factor (TNF)-α, TNF-β, and IL-1 receptor antagonist. In addition, analysis of annotations revealed that this PPI network was enriched in immune functions, such as “defense response”, “inflammatory response”, “cytokine-mediated signaling pathway”, and “chemotaxis” [30]. We also identified 5 downregulated genes, namely IL-10, CCL3, colony-stimulating factor (CSF)1, vascular endothelial growth factor (VEGF), and NGF (neuronal growth factor) and extracted one validated PC (labeled: PCdown) [30]. Further analysis revealed that the PPI network of these downregulated genes was enriched in functions such as “positive regulation of neurogenesis”, “regulation of neuron death”, “negative regulation of neural death”, and “regulation of microglial cell migration”. Consequently, we have constructed a new composite which reflects immune-inflammatory pathways versus lowered regulation of key neuroprotective functions as z immune network index – z PCdown (labeled as immune-linked neurotoxicity index or INT).

As explained previously [7,18], the concentrations of total cholesterol, HDL-cholesterol, triglycerides, and direct LDL-cholesterol were determined using the Alinity C (Abbott Laboratories, USA; Otawara-Shi, Tochigi-Ken, Japan). The ApoA1 and ApoB concentrations were determined using immunoturbidimetric assays with the Roche Cobas 6000 and c501 modules from Roche (Rotkreuz, Switzerland). By utilizing the Free Cholesterol Colorimetric Assay reagent (Elabscience, cat number: E-BC-K004-M) we measured free cholesterol. Consequently, we constructed a new atherogenic index as z ApoB + z triglycerides + z free cholesterol + z LDL-cholesterol – z HDL-cholesterol - z (1-free cholesterol / total cholesterol) – z ApoA1 [18]. Finally, a new biomarker index that integrated increased INT and atherogenicity was constructed as z INT index + z atherogenicity index (labeled as METAMMUNE). As such, in the current study we used three biomarker indices: immune network index, INT, and METAMMUNE. Since previously [18] we observed significant interactions between MetS and immune network index we also included this interaction term and the ROI x MetS interaction.

### Statistical analysis

To achieve feature reduction, PCA was conducted to consolidate the variables into a singular PC score. For determining factorability, we employed the Bartlett’s sphericity test and the Kaiser-Meyer-Olkin test for sample adequacy. To create clinical PC models that accurately represent an underlying concept, specific criteria were used: the first PC was deemed acceptable if it explained more than 50% of the variance, all loadings on the initial PC were above 0.66, and Cronbach’s alpha exceeded 0.7.

An analysis of variance (ANOVA) was used to examine scale variables across different diagnostic groups. For the purpose of comparing nominal variables across different categories, we employed either the chi-square test or Fisher’s Exact Probability Test. To establish the connections between two sets of scale variables, we utilized either Pearson’s product moment or Spearman’s rank order correlation coefficients. A thorough analysis was conducted to examine the connections between various explanatory variables (ACEs, biomarkers) and dependent variables (clinical phenome data) through the utilization of multiple regression analysis. We employed a manual approach and further utilized a forward stepwise automatic regression method to determine the most accurate predictors for the model. We set inclusion and exclusion p-values at 0.05 and 0.06, respectively. The final regression models included standardized coefficients, t-statistics, and exact p-values for each significant explanatory variable. Additionally, the total variance was represented by effect size measures such as partial eta squared (R^2^), or and F statistics (along with p values) were also considered. The presence of heteroskedasticity was assessed using the White and modified Breusch-Pagan test. The analysis of collinearity and multicollinearity involved the examination of various factors such as tolerance (with a cut-off value of less than 0.25), variance inflation factor (with a cut-off value of greater than 4), condition index, and variance proportions from the collinearity diagnostics table.

The data distributions were subjected, where needed, to various normalization processes, such as rank-based, logarithmic, square-root, and inverse normal transformations. Furthermore, the data underwent z-score transformation to enhance their interpretability and generate z-unit-weighted composite scores that accurately depict distinct immune profiles. In each of the investigations mentioned, a two-tailed design was used to determine statistical significance at an α value of 0.05. The software used was IBM Windows SPSS version 29.

The estimated a priori sample size was calculated using G*Power 3.1.9.4 and applied to the primary analysis, which involved conducting a multiple regression analysis of ROI on ACEs and biomarkers. Based on an effect size f value of 0.25 (which accounts for approximately 10% of the variance), along with a maximum of 5 explanatory variables, an alpha value of 0.05, and a power of 0.8, it was determined that a minimum sample size of 122 was necessary. It should be added that the post-hoc estimated power for the same analysis was 0.999.

## Results

### Sociodemographic, clinical and biomarker data in patients and controls

**Table 1** shows the sociodemographic and clinical data of the patients and controls in the current study. There were no significant differences in age, sex ratio, education years, marital status, current smoking, BMI, MetS ratio, mild COVID-19 infection, between the two study groups. The unemployment rate was significantly higher in OMDD patients than in controls. All clinical scores were significantly higher in patients than in controls. This table also displays the mean values of the three biomarkers in patients versus controls. The three biomarker scores were significantly increased in patients as compared with controls.

**Table 1.**
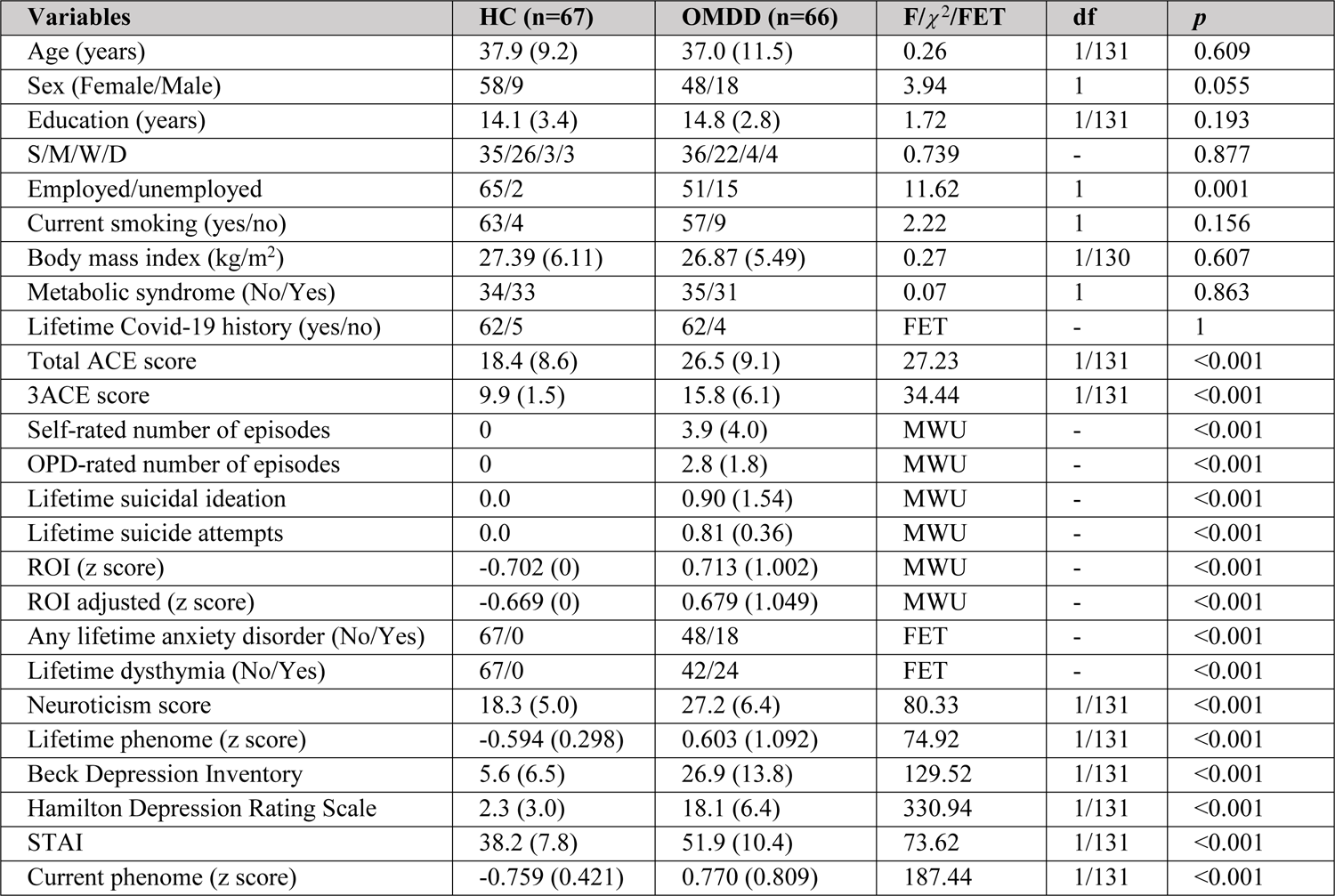
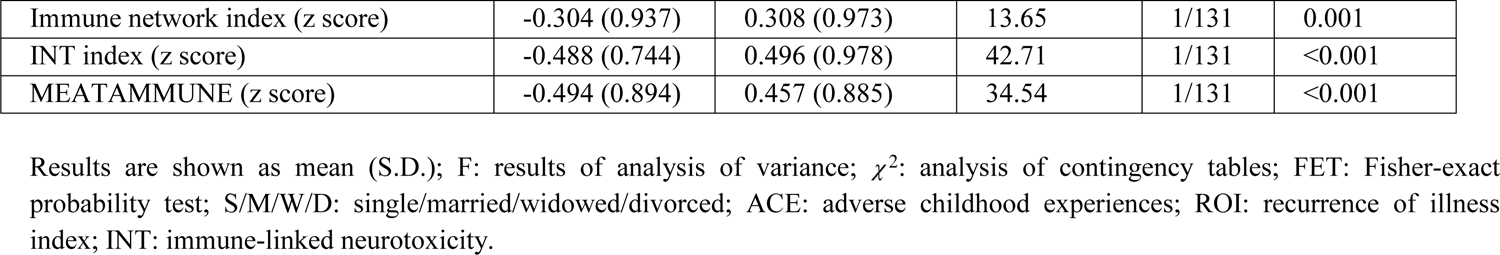
Demographic and clinical data of outpatients with major depressive disorder (OMDD) and healthy controls (HC)

| Variables | HC (n=67) | OMDD (n=66) | F/$\chi^2$/FET | df | p |
| --- | --- | --- | --- | --- | --- |
| Age (years) | 37.9 (9.2) | 37.0 (11.5) | 0.26 | 1/131 | 0.609 |
| Sex (Female/Male) | 58/9 | 48/18 | 3.94 | 1 | 0.055 |
| Education (years) | 14.1 (3.4) | 14.8 (2.8) | 1.72 | 1/131 | 0.193 |
| S/M/W/D | 35/26/3/3 | 36/22/4/4 | 0.739 | - | 0.877 |
| Employed/unemployed | 65/2 | 51/15 | 11.62 | 1 | 0.001 |
| Current smoking (yes/no) | 63/4 | 57/9 | 2.22 | 1 | 0.156 |
| Body mass index (kg/m<sup>2</sup>) | 27.39 (6.11) | 26.87 (5.49) | 0.27 | 1/130 | 0.607 |
| Metabolic syndrome (No/Yes) | 34/33 | 35/31 | 0.07 | 1 | 0.863 |
| Lifetime Covid-19 history (yes/no) | 62/5 | 62/4 | FET | - | 1 |
| Total ACE score | 18.4 (8.6) | 26.5 (9.1) | 27.23 | 1/131 | <0.001 |
| 3ACE score | 9.9 (1.5) | 15.8 (6.1) | 34.44 | 1/131 | <0.001 |
| Self-rated number of episodes | 0 | 3.9 (4.0) | MWU | - | <0.001 |
| OPD-rated number of episodes | 0 | 2.8 (1.8) | MWU | - | <0.001 |
| Lifetime suicidal ideation | 0.0 | 0.90 (1.54) | MWU | - | <0.001 |
| Lifetime suicide attempts | 0.0 | 0.81 (0.36) | MWU | - | <0.001 |
| ROI (z score) | -0.702 (0) | 0.713 (1.002) | MWU | - | <0.001 |
| ROI adjusted (z score) | -0.669 (0) | 0.679 (1.049) | MWU | - | <0.001 |
| Any lifetime anxiety disorder (No/Yes) | 67/0 | 48/18 | FET | - | <0.001 |
| Lifetime dysthymia (No/Yes) | 67/0 | 42/24 | FET | - | <0.001 |
| Neuroticism score | 18.3 (5.0) | 27.2 (6.4) | 80.33 | 1/131 | <0.001 |
| Lifetime phenome (z score) | -0.594 (0.298) | 0.603 (1.092) | 74.92 | 1/131 | <0.001 |
| Beck Depression Inventory | 5.6 (6.5) | 26.9 (13.8) | 129.52 | 1/131 | <0.001 |
| Hamilton Depression Rating Scale | 2.3 (3.0) | 18.1 (6.4) | 330.94 | 1/131 | <0.001 |
| STAI | 38.2 (7.8) | 51.9 (10.4) | 73.62 | 1/131 | <0.001 |
| Current phenome (z score) | -0.759 (0.421) | 0.770 (0.809) | 187.44 | 1/131 | <0.001 |
| Immune network index (z score) | -0.304 (0.937) | 0.308 (0.973) | 13.65 | 1/131 | 0.001 |
| INT index (z score) | -0.488 (0.744) | 0.496 (0.978) | 42.71 | 1/131 | <0.001 |
| MEATAMMUNE (z score) | -0.494 (0.894) | 0.457 (0.885) | 34.54 | 1/131 | <0.001 |

Results are shown as mean (S.D.); F: results of analysis of variance; $\chi^2$ : analysis of contingency tables; FET: Fisher-exact probability test; S/M/W/D: single/married/widowed/divorced; ACE: adverse childhood experiences; ROI: recurrence of illness index; INT: immune-linked neurotoxicity.

### Number of episodes and construction of the ROI scores

There were significant associations between the OPD-rated and self-rated number of episodes, using Pearson’s (r=0.804, p<0.001, n=133) and Spearman’s rank order (r=0.981, p<0.001) correlation coefficients. In the restricted group of OMDD patients, there were significant associations between the OPD-rated and self-rated number of episodes using Pearson’s (r=0.690, p<0.001, n=66) and Spearman’s rank order (r=0.862, p<0.001) correlation coefficients. Using the Wilcoxon Signed Rank test, the median of differences between OPD-rated and self-rated episode number was significantly different (p<0.001). Table 1 shows that the number of depressions was significantly higher when self-rated as compared with the clinician OPD ratings.

**Table 2** shows how we constructed the first ROI index based on PC analysis performed on number of OPD-rated episodes (in log10 transformation), LT_SI and LT-SA. The first PC explained 75.02% of the total variance and Cronbach’s alpha was 0.819. All three variables loaded highly on this PC (>0.801). We also performed a PCA on the adjusted OPD-rated number of episodes, and both LT_SB data and found that the first PC explained 74.61% of the variance, all loadings were > 0.795 and Cronbach’s alpha was more than adequate at 0.815 (labeled adjusted ROI). As such, two reliable PCs reflecting ROI could be constructed.

**Table 2.** Results of principal component analysis.

| ROI |  | ROI pathtway phenotype |  |
| --- | --- | --- | --- |
| Variables | Loading | Variables | Loading |
| Number of episodes | 0.801 | Number of episodes | 0.778 |
| Lifetime suicidal ideation | 0.934 | Lifetime suicidal ideation | 0.832 |
| Lifetime suicidal attempts | 0.858 | Lifetime suicidal attempts | 0.734 |
|  |  | INT index | 0.716 |
|  |  | METAMMUNE index | 0.737 |
| KMO = 0.622 |  | KMO = 0.663 |  |
| X<sup>2</sup> = 181.793 (df=3), p<0.001 |  | X<sup>2</sup> = 331.921 (df=10), p<0.001 |  |
| VE = 75.02% |  | VE = 57.82% |  |
| Cronbach’s alpha=0.819 |  | Cronbach’s alpha=0.810 |  |

KMO: The Kaiser-Meyer-Olkin Test; VE: Variance explained; $X^2$ : Bartlett's test of sphericity.
INT: immune-linked neurotoxicity index

### ACEs predict ROI

**Table 3** shows the results of multiple regression analyses with the ROI scores as dependent variable and ACEs as explanatory variables. We found that (regression #1) 19.1% of the variance in the OPD-rated number of episodes was explained by total sum of ACEs, and that 22.7% of the variance in ROI (regression #2) was explained by total ACE score and male sex. The same variables explained 22.6% of the variance in the adjusted ROI score (F=19.02, df=2/1030, p<0.001; total AEC score: t=5.82, p<0.001; male sex: t=2.21, p=0.029). To define the most important types of ACEs, the total sum ACE score was deleted from the analysis, indicating that divorce, physical and sexual abuse were the most prominent ACE subtypes (regression #3). **Figure 1** shows the partial regression of ROI on the 3ACEs score (after adjusting for age, sex, education). Consequently, we have entered the three biomarker scores in the analysis and detected that 30.5% of the variance in ROI was explained (regression #4) by divorce, emotional abuse and neglect and the METAMMUNE score.

**Figure 1.**
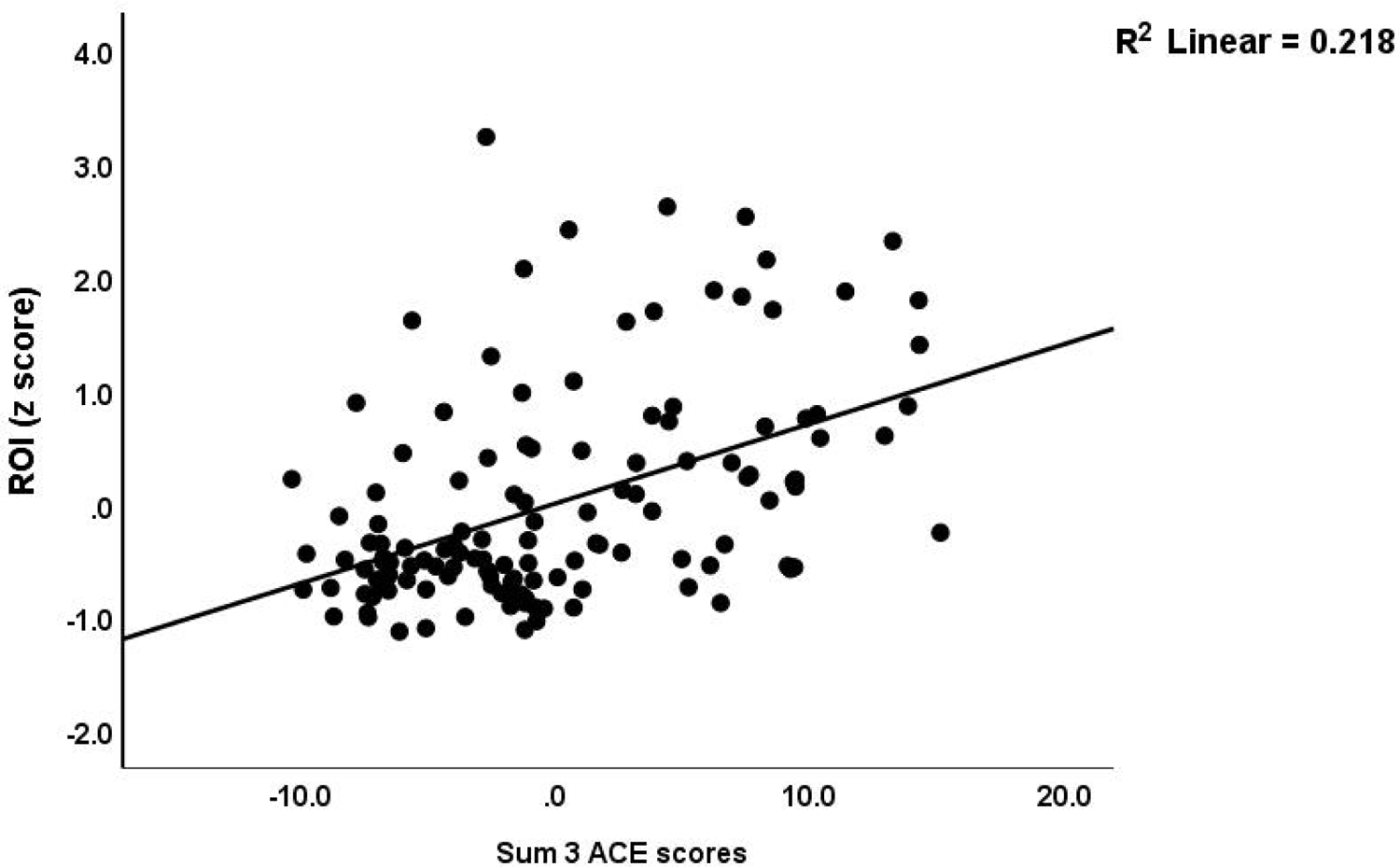
Partial regression of the recurrence of illness (ROI) index on the sum of three selected adverse childhood experience (ACE) domains (p<0.01)

**Table 3.**
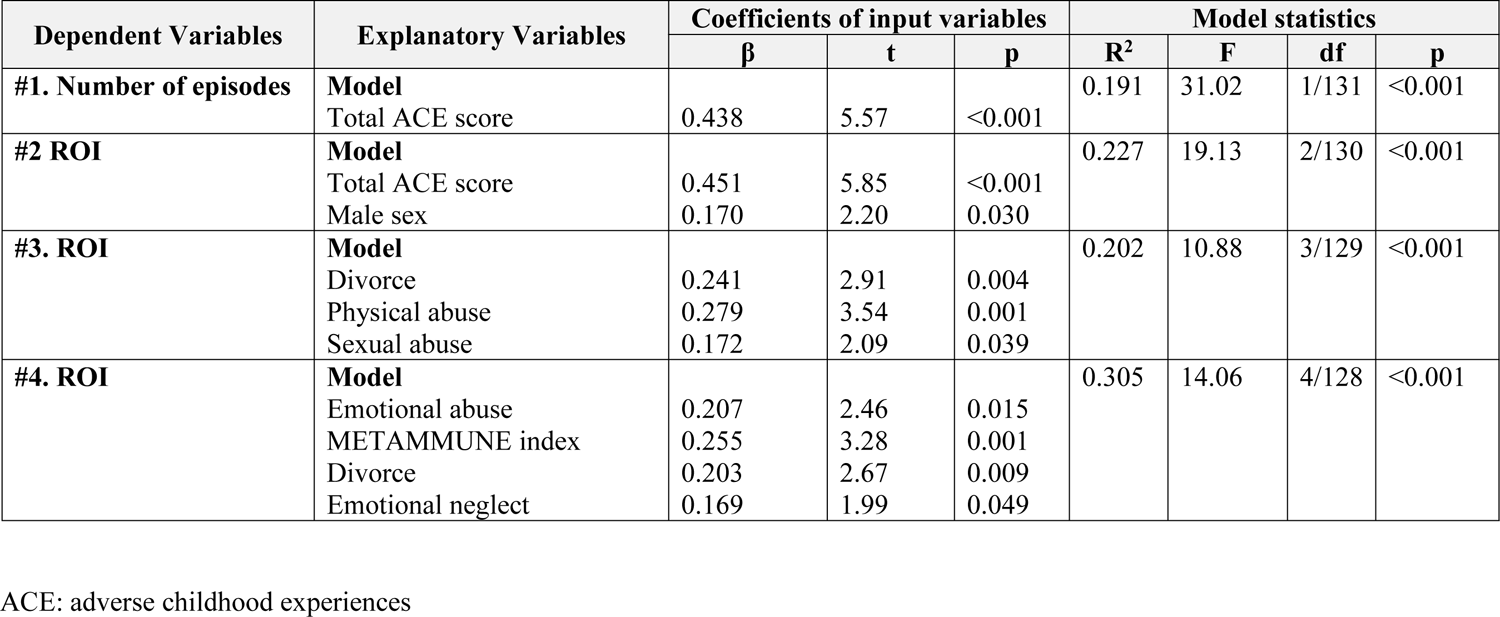
Results of multiple regression analyses with recurrence of illness (ROI) data as dependent variables and adverse childhood experiences and biomarkers as explanatory variables.

| Dependent Variables | Explanatory Variables | Coefficients of input variables |  |  | Model statistics |  |  |  |
| --- | --- | --- | --- | --- | --- | --- | --- | --- |
| | | $\beta$ | t | p | R<sup>2</sup> | F | df | p |
| #1. Number of episodes | **Model**<br>Total ACE score | 0.438 | 5.57 | <0.001 | 0.191 | 31.02 | 1/131 | <0.001 |
| #2 ROI | **Model**<br>Total ACE score | 0.451 | 5.85 | <0.001 | 0.227 | 19.13 | 2/130 | <0.001 |
|  | Male sex | 0.170 | 2.20 | 0.030 |  |  |  |  |
| #3. ROI | **Model**<br>Divorce | 0.241 | 2.91 | 0.004 | 0.202 | 10.88 | 3/129 | <0.001 |
|  | Physical abuse | 0.279 | 3.54 | 0.001 |  |  |  |  |
|  | Sexual abuse | 0.172 | 2.09 | 0.039 |  |  |  |  |
| #4. ROI | **Model**<br>Emotional abuse | 0.207 | 2.46 | 0.015 | 0.305 | 14.06 | 4/128 | <0.001 |
|  | METAMMUNE index | 0.255 | 3.28 | 0.001 |  |  |  |  |
|  | Divorce | 0.203 | 2.67 | 0.009 |  |  |  |  |
|  | Emotional neglect | 0.169 | 1.99 | 0.049 |  |  |  |  |

ACE: adverse childhood experiences

### Correlations between ROI and clinical data

**Table 4** shows the correlations between the ROI score and all clinical data in the total study group and in the restricted study sample of OMDD patients. In the former study sample, ROI was significantly correlated with all clinical data including ACE scores, severity of the LT and current phenome and LT and current SBs and the biomarker data as well. **Figures 2** and **3** show the partial regressions of the METAMUNNE and INT indices on ROI scores (after adjusting for sex, age, BMI, and ACEs). In subjects with OMDD, ROI was significantly correlated with the ACE scores, neuroticism, LT and current phenome, SB, and the METAMMUNE index.

**Figure 2.**
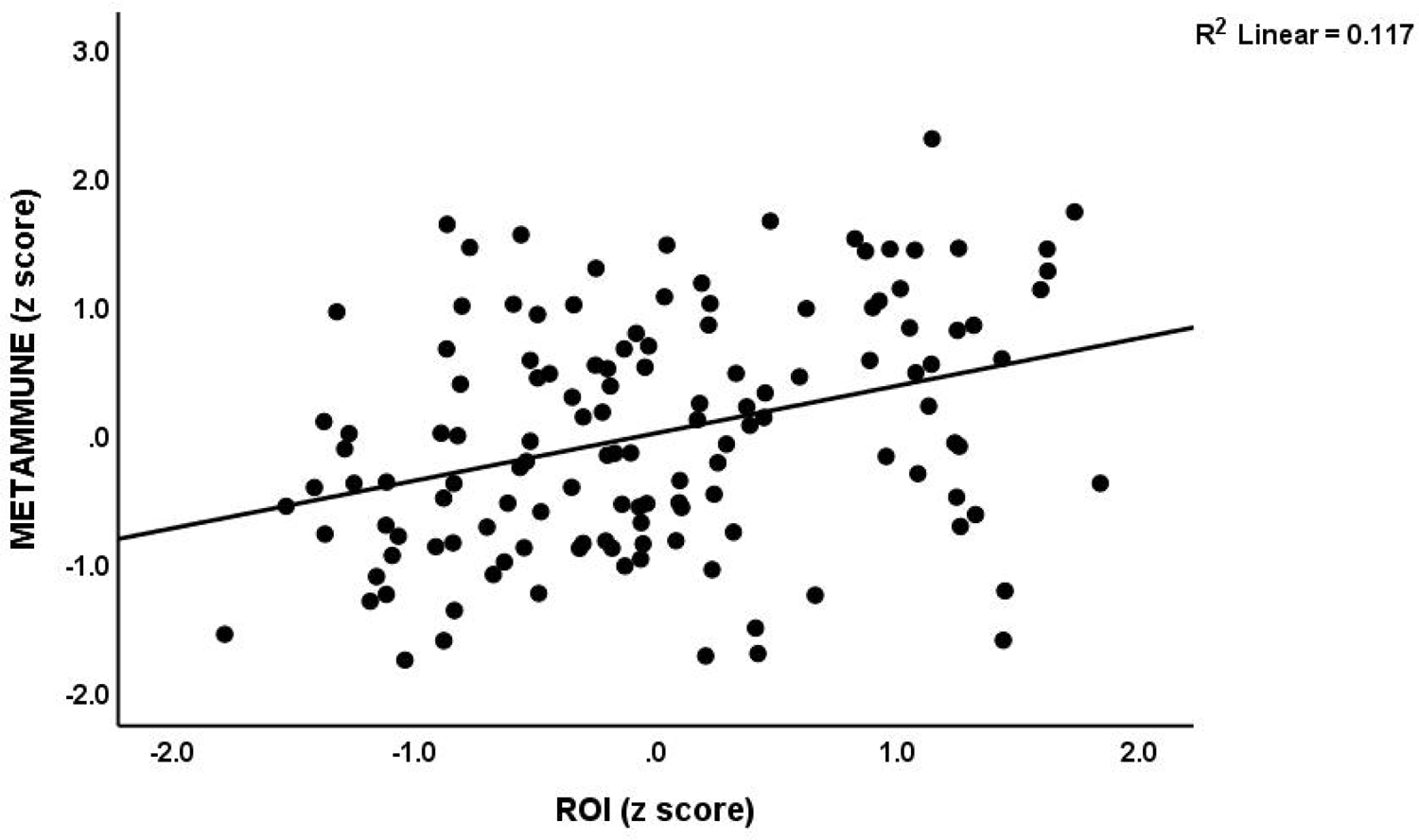
Partial regression of the METAMMUNE index on recurrence of illness (ROI) scores (p<0.01).

**Figure 3.**
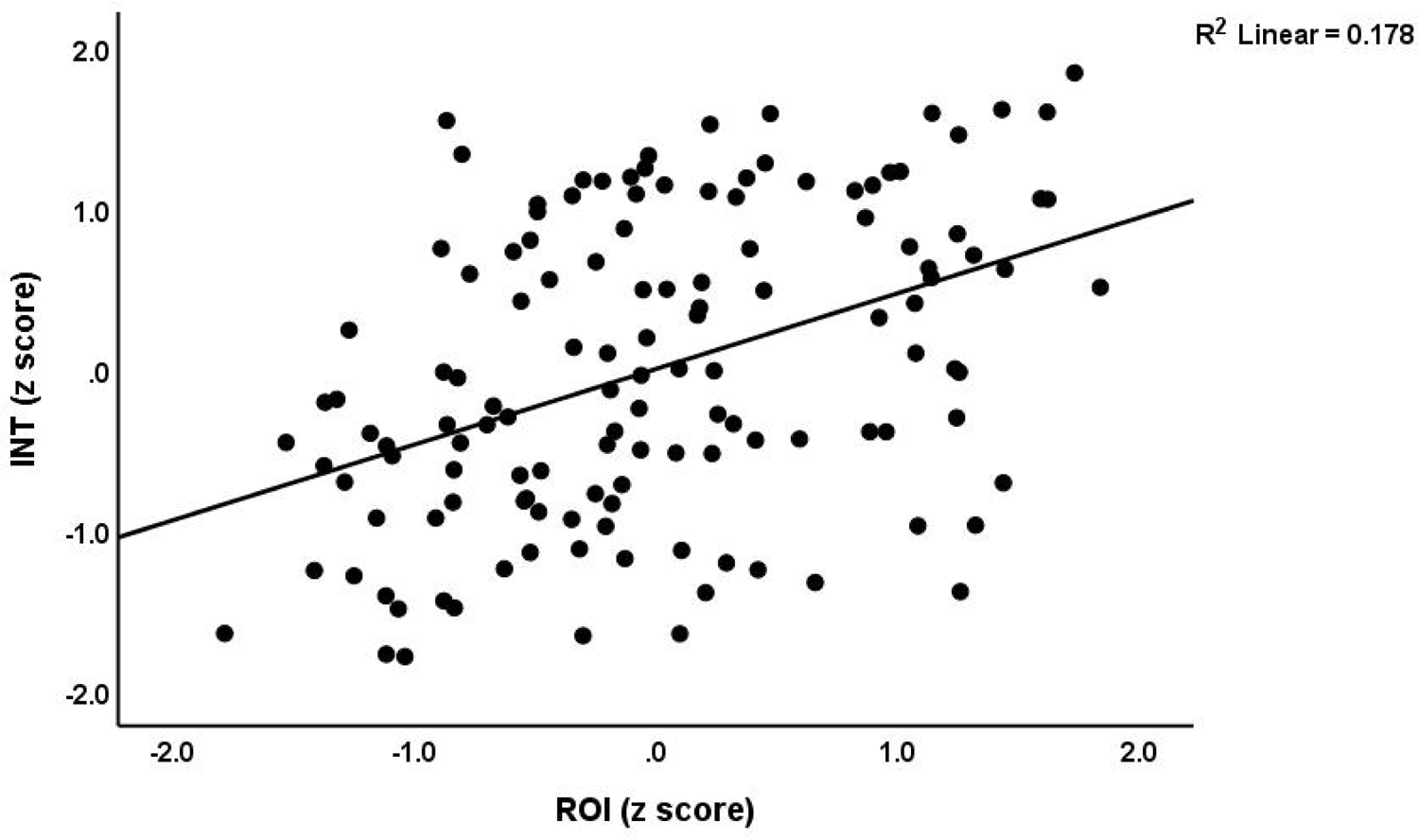
Partial regression of the INT (immune-linked neurotoxicity) index on recurrence of illness (ROI) scores (p<0.01).

**Table 4.** Correlation matrix between recurrence of illness (ROI) index and clinical and biomarker data.

| **Variables** | **ROI in all subjects**<br>**N=133** | **ROI in MDD only**<br>**N=66** |
| --- | --- | --- |
| 3ACE score | 0.503 ** | 0.254 * |
| Total ACE score | 0.509 ** | 0.274 * |
| Neuroticism score | 0.640 ** | 0.312 * |
| Lifetime phenome | 0.714 ** | 0.414 * |
| Hamiton Depression Rating Scale score | 0.823 ** | 0.209 |
| Beck Depression Inventory score | 0.732 ** | 0.318 * |
| State Train Anxiety Inventory, state score | 0.603 ** | 0.206 |
| Current suicidal behaviours | 0.689 ** | 0.486 ** |
| Current phenome | 0.764 ** | 0.312 * |
| Immune network index | 0.328 ** | 0.094 |
| Immune-linked neurotoxicity index | 0.461 ** | 0.049 |
| METAMMUNE INDEX | 0.336 ** | 0.256 * |

Results of Spearman's rank-order correlation calculations; \*\*p<0.01; \* p<0.05.

**Table 5** shows the results of multiple regression analyses with the clinical outcome data as dependent variables and ACEs scores and biomarkers as explanatory variables. Regression #1 shows that 47.4% of the variance in the LT_phenome was associated with ROI and emotional abuse. **Figure 4** shows the partial regression of the lifetime phenome score on ROI (after controlling for sex, education, BMI, and MetS). Regression #2 shows that 55.1% of the variance in the current_phenome was explained by ROI, INT index, emotional neglect and abuse. **Figure 5** shows the partial regression of the current phenome on ROI after adjusting for sex, age, education, body mass index, and MetS). In the restricted OMDD group (regression #3), we found that 40.9% of the variance in the current_phenome was explained by ACEs (sexual abuse and emotional neglect) and the interaction term ROI x MetS.

**Figure 4.**
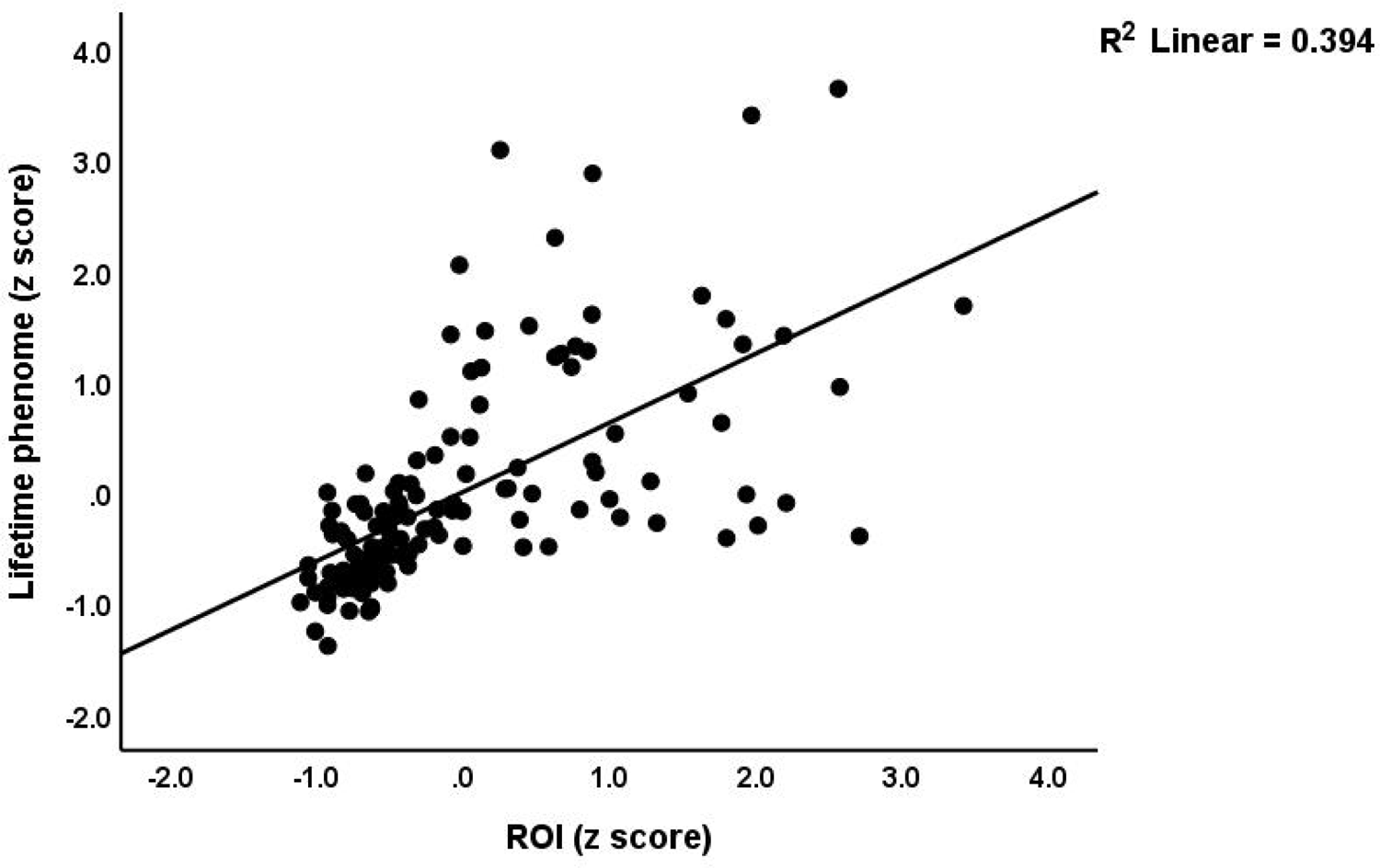
Partial regression of the lifetime phenome score on the recurrence of illness (ROI) index (p<0.01)

**Figure 5.**
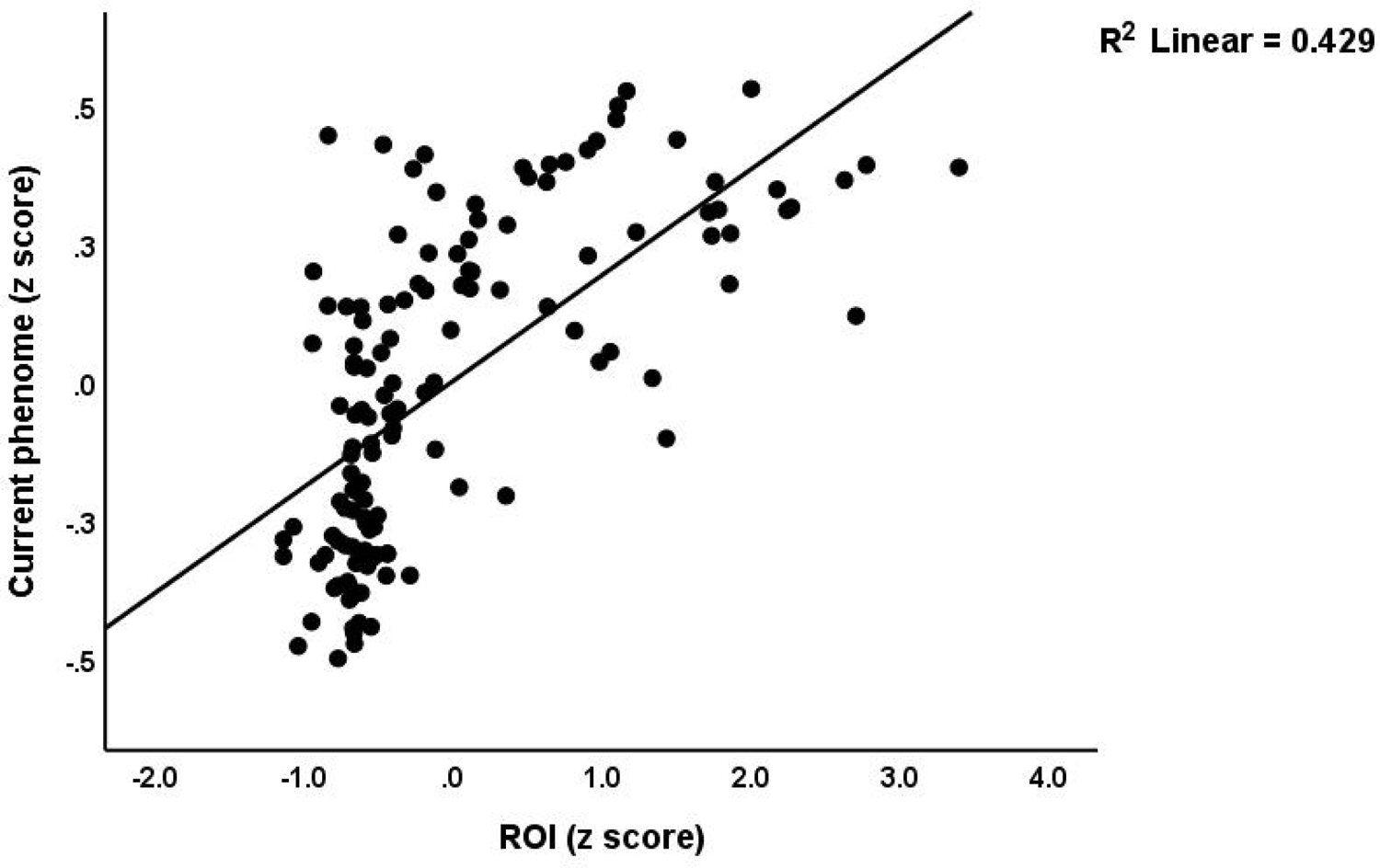
Partial regression of the current phenome on the recurrence of illness (ROI) index (p<0.01)

**Table 5.** Results of multiple regression analyses with clinical scores as dependent variables and biomarkers and adverse childhood experiences as explanatory variables.

| Dependent Variables | Explanatory Variables | Coefficients of input variables |  |  | Model statistics |  |  |  |
| --- | --- | --- | --- | --- | --- | --- | --- | --- |
| | | $\beta$ | t | p | R<sup>2</sup> | F | df | p |
| #1. LT_phenome | **Model** |  |  |  | 0.474 | 58.49 | 2/130 | <0.001 |
|  | ROI | 0.549 | 7.94 | <0.001 |  |  |  |  |
|  | Emotional abuse | 0.253 | 3.67 | <0.001 |  |  |  |  |
| #2. Current_phenome | **Model** |  |  |  | 0.551 | 39.29 | 4/128 | <0.001 |
|  | ROI | 0.444 | 6.42 | <0.001 |  |  |  |  |
|  | Emotional neglect | 0.188 | 2.72 | 0.007 |  |  |  |  |
|  | INT | 0.209 | 3.26 | 0.001 |  |  |  |  |
|  | Emotional abuse | 0.164 | 2.38 | 0.019 |  |  |  |  |
| #3. Current_phenome in MDD | **Model** |  |  |  | 0.409 | 10.57 | 4/61 | <0.001 |
|  | Age | -0.398 | -3.98 | <0.001 |  |  |  |  |
|  | Emotional neglect | 0.355 | 3.50 | 0.001 |  |  |  |  |
|  | Sexual abuse | 0.228 | 2.24 | 0.029 |  |  |  |  |
|  | ROI x MetS | 0.221 | 2.23 | 0.029 |  |  |  |  |

ROI: recurrence of illness index; INT: immune-linked neurotoxicity; MetS: metabolic syndrome

The associations between the ROI components and the INT and METAMMUNE indices were so important that one validated PC could be extracted from the 5 variables. The construction of this PC is shown in Table 2 (labeled ROI-pathway phenotype). Cronbach’s alpha was 0.810.

## Discussion

### Computation of ROI

A first notable finding of this study is that individuals’ self-rated number of episodes aligns reasonably well with the actual number of episodes determined by physicians. This is evident from the significant Spearman correlation coefficient between the self-rated and physician-rated number of episodes. However, the number of episodes that patients reported was higher than what physicians rated, indicating that patients may have somewhat exaggerated their own number of episodes. However, the disparities are reduced once the self-rated data is transformed using transformations (e.g. rank order). Therefore, the self-rated number of episodes are valuable for calculating the ROI index albeit they may have to be processed in transformations.

In the current study focusing on Thai OMDD, a ROI index was successfully developed using the number of episodes (in log or rank transformations) and the frequency of lifetime suicidal ideation and suicidal behaviors. In previous studies, similar ROI indices were computed for OMDD patients in Brazil and Thailand [3,4,5,6,7,9]. It is crucial to understand that these factors must demonstrate high quality fit standards, such as having satisfactory AVE, Cronbach’s α or composite reliability values, and displaying appropriate loadings (>0.66) for all variables on the latent vectors.

Furthermore, it is worth noting that researchers have also developed a comparable ROI index for individuals diagnosed with bipolar disorder (BD) [3,4,31]. From the number of depressive episodes, (hypo)manic episodes, and frequency of suicidal ideation and self-harm, researchers have identified a consistent factor in BP patients [3,4,31]. These results show that the recurrence of depressive, (hypo)manic episodes, and suicidal behaviors are manifestations of the same underlying concept, namely ROI.

It is widely known that mood disorders, such as MDD and BD, are recurrent disorders with a higher risk for suicidal behaviors, including suicide attempts [32–34]. Nevertheless, course trajectory specifiers have been largely overlooked in the diagnostic systems, including those of the DSM and ICD. Disorders in the DSM/ICD are categorized according to their symptoms and symptom clusters, along with few course specifiers like bipolarity and rapid cycling. A recent project proposed the development of a clinical course-graphing scale for DSM-5 disorders, which is an interesting concept to consider. They introduced the Timeline Course Graphing Scale for the DSM-5 Mood Disorders (TCGS) [35], which is a specific tool used for tracking the progression of mood disorders. This approach utilizes a meticulously organized technique for mapping out the course of mood disorders, allowing researchers to make well-informed evaluations regarding the timing of onset (early or late) and the severity of the illness (chronicity, subthreshold syndrome, etc.). Unfortunately, the main goal of the TCGS is to distinguish between MDD and the new DSM-5 classification called Persistent Depression, rather than measuring ROI scores. A different approach to examining the alternating symptoms in mood disorders is the NIMH Life Chart Method (LCM-p) [36]. Earlier studies conducted by Kapczinski et al. [11], Berk et al. [10], and Ferensztajn et al. [12] put forth staging indices that were developed using a theoretical framework. However, all these staging indices lacked validated mathematical ROI algorithms as the ROI constructed in the current study.

### Connections between ACEs and ROI

Through this study, it has been determined that the cumulative effect of ACEs such as parental divorce, physical abuse, and sexual abuse, have a significant impact on the variance in ROI, accounting for approximately 20-22.7% of the variance. In previous studies, it has been found that ACEs highly predicted ROI in individuals with OMDD and bipolar disorder (BD) in Brazil, as well as in patients with severe OMDD in Thailand [1,3,4,5,7,18]. In addition, a recent study revealed a significant correlation between a risk/resilience ratio and the ROI score. The researchers computed this risk/resilience ratio by dividing the ACE sum score by a composite of antioxidant defenses, namely an antioxidant gene (Q192R paraoxonase 1 gene variant and its corresponding enzyme PON1). Remarkably, this ratio accounted for a substantial 20.4% of the variance in the ROI score [4].

In the present study, it was observed that there was a strong and significant correlation between ROI and the lifetime phenome, which is a composite of neuroticism, dysthymia, and lifetime anxiety disorders. Additionally, ROI was found to be significantly associated with the current phenome. In previous studies, it was reported that the combined impacts of different ACE factors, which influence outcome variables like the current phenome and current SBs, are partially mediated by ROI [1,3,4,5] and that ROI, in turn, is correlated with the presence of functional disabilities, neurocognitive deficits, and progressive deterioration [1,3]. These results extend previous studies which have shown a correlation between ACEs and a higher frequency of recurring episodes [37,38,39], elevated risk for subsequent suicidal behaviors [40,41], more severe depression [42–46], and a decline in HR-QoL [47–50].

### Associations between ROI and INT biomarkers

The major finding of the current study is the strong correlation between ROI and biomarkers of an activated immune network, INT, and the interaction between INT and metabolic disorders including atherogenicity. Previous studies have demonstrated a correlation between a higher frequency of episodes and relapses with pro-inflammatory cytokines, C-reactive protein, neopterin, and oxidative stress damage, as indicated by malondialdehyde [51,52,53]. Earlier research has demonstrated a correlation between ROI and a higher body mass index (BMI), the metabolic syndrome (MetS), and elevated insulin resistance [54,55]. More recent studies have shown that ROI is associated with cytokine networks, T cell activation and lowered T regulatory cells, oxidative damage, lowered antioxidant and neurotrophic factors, and a specific gut dysbiosis endotype [5,6,7,8,56,57,58].

In addition, the study successfully extracted a reliable PC from the three ROI components along with the INT and METAMMUNE indices. This suggests that both ROI and these biomarkers are expressions of the same latent concept, which is a ROI-neuroimmune-metabolic pathway phenotype. In a previous discussion, we highlighted the potential of the precision nomothetic approach in creating new pathway phenotypes [1]. This involves effectively combining phenotypic features with biomarker pathways in replicable and validated latent vectors [1]. In previous studies, pathway phenotypes in MDD have been identified, such as the construction of a ROI-neuroimmune-growth factor pathway phenotype [5]. In a separate study, the first author’s laboratory successfully developed a pathway phenotype by utilizing ROI and paraoxonase 1 (PON1) gene variants along with PON1 enzymatic activity which ultimately increased neuro-oxidative neurotoxicity [1,3]. Thus, the combination of ROI and immune or oxidative neurotoxic pathways plays a crucial role in shaping the phenome of MDD. Importantly, there is evidence that episodes of MDD and (hypo)mania can have negative impacts on neuronal functions in the brain as demonstrated by changes in neurocognitive functions, overall psychological disabilities, and brain imaging techniques [59]. Therefore, when ACEs are combined with reduced protective factors (including genetically determined antioxidant levels), it can lead to a cycle where the impact on the brain’s reward system and toxic pathways becomes more pronounced, ultimately increasing the risk and severity of the current phenome.

In the context of temporal lobe epilepsy (TLE), an intriguing observation emerges. A pathway phenotype has been constructed by considering various symptoms of general psychopathology, such as depression and anxiety, along with factors like oxidative stress toxicity and reduced antioxidant levels [60]. In addition, the researchers found that the frequency of episodes in TLE was linked to higher levels of neurotoxicity, as indicated by increased neuro-oxidative stress and reduced antioxidant defenses, including PON1 activity [60]. These recent findings indicate a progressive rise in “epileptogenic kindling” caused by the combined impact of neurotoxicity. As such, the “kindling” of episode frequency in epilepsy and ROI in MDD are both linked to neurotoxicity processes.

### Clinical relevance of ROI

From the information provided, it can be inferred that ROI plays a significant role in mood disorders. Increasing ROI suggests a repeated activation of the immune system and oxidative stress and the resulting neurotoxicity leading to higher susceptibility to a relapsing-remitting pattern, more severe lifetime symptoms such as neuroticism, dysthymia, and anxiety disorders, as well as increased severity of new episodes and suicidal behaviors. Moreover, ROI plays a crucial role in mediating the effects of gene variants including the PON1 Q192R gene and ACEs on the phenome [3] and, therefore, determines the lifelong trajectory of individuals with MDD, starting from genetic variants and ACEs, leading to the development of behavioral deficits.

From a clinical perspective, it is important to consider the ROI index when assessing the patient’s condition and, therefore, ROI should be incorporated into the clinical diagnosis. In a recent study, a novel clinimetrics method called Research and Diagnostic Algorithmic Rules (RADAR) was developed which encompasses quantitative scores of all relevant features of MDD, including ACEs, ROI, and phenome features [6]. Moreover, these quantitative RADAR scores may be presented graphically in RADAR or spider graphs [6,31]. These graphs provide a visual representation of all patient’s characteristics, including ROI, thus shaping a unique fingerprint. By simply looking at the spider graphs, one can quickly estimate the key traits and severity of the illness. Therefore, it is important for biomarker research in mood disorders to utilize these RADAR scores to analyze associations between biomarker pan-omics data and the relevant clinical scores, including ROI, lifetime and current suicidal behaviors and phenome characteristics. Hence, it is more beneficial to employ our quantitative scores of ROI and the phenome in statistical analyses or machine learning techniques rather than depending on incorrect binary DSM/ICD diagnoses [1,6,31].

### Limitations

Future research should investigate the effects of ROI on brain imaging data including metabolic and hemodynamic changes as measured with functional near infrared spectroscopy, the brain connectome as measured with functional magnetic resonance imaging [61,62], and neuronal damage markers of depression [63]. Since the studies of the first author were performed in Thai and Brazil patients, the results deserve replication in other countries and ethnicities. Lastly, more research is needed on the impact of ROI in MDD versus BD patients [31].

## Conclusions

This study constructed a ROI index based on number of physician-rated depressive episodes, frequency of suicidal ideation and attempts by extracting one validated principal component. Around 20-22% of the variance in this ROI score was explained by ACEs. ROI was significantly associated with the INT and METAMMUNE indices, neuroticism, lifetime and current suicidal behaviors, and the severity of the OMDD phenome. We found that 55.1% of the variance in the phenome of OMDD is explained by INT, ROI, and ACEs. We were able to extract a validated latent construct from the three ROI components, INT and METAMMUNE indices, indicating that ROI and the immune-metabolic pathways are expressions of the same latent concept, namely a ROI-neuroimmune-metabolic pathway phenotype.

All in all, ROI is a key factor indicating increasing immune-metabolic aberrations and vulnerability to develop new episodes and suicidal behaviors. Future research should always ROI-define the study samples and examine the associations between ROI and biomarkers, and the mediating effects of ROI on the depression phenome. Consequently, ROI is a new drug target to treat depression and prevent further episodes and suicidal behaviors. Consequently, novel treatments should target the immune-linked neurotoxicity pathways and atherogenicity delineated in OMDD.

### Ethics approval and consent to participate

The research project (#445/63) was approved by the Institutional Review Board of Chulalongkorn University’s institutional ethics board. All patients and controls gave written informed consent prior to participation in the study.

## Author’s contributions

Conceptualization and study design: MM and JK; first draft writing: MM; statistical analysis: MM; editing: all authors; recruitment of patients: JK. All authors approved the last version of the manuscript.

## Funding

This work was supported by the Ratchadapiseksompotch Fund, Graduate Affairs, Faculty of Medicine, Chulalongkorn University (Grant number GA64/21), a grant from CU Graduate School Thesis Grant, and Chulalongkorn University Graduate Scholarship to Commemorate the 72nd Anniversary of His Majesty King Bhumibol Adulyadej to KJ; the Thailand Science research, and Innovation Fund Chulalongkorn University (HEA663000016), and a Sompoch Endowment Fund (Faculty of Medicine) MDCU (RA66/016) to MM, and Grant No BG-RRP-2.004-0007-С01, Strategic Research and Innovation Program for the Development of MU - PLOVDIV - (SRIPD-MUP), Creation of a network of research higher schools, National plan for recovery and sustainability, European Union - NextGenerationEU.

## Conflict of interest

The authors have no commercial or other competing interests concerning the submitted paper.

## Data Availability

The dataset generated during and/or analyzed during the current study will be available from the corresponding author upon reasonable request and once the dataset has been fully exploited by the authors.

## Notes

### Competing Interest Statement

The authors have declared no competing interest.

